# Using comprehensive disease modeling to assess the burden of substandard and falsified oxytocin in Kenya

**DOI:** 10.1101/2023.05.04.23289537

**Authors:** Sabra Zaraa, Josh J Carlson, Elisabeth Vodicka, Andy Stergachis

**Author notes:** Corresponding author. (206)468-9262.

## Abstract

**BACKGROUND:** Substandard and falsified (SF) oxytocin threatens the health of pregnant patients, resulting in prolonged illness and severe avertable disease outcomes. Additionally, SF oxytocin leads to an economic burden on the healthcare system and society due to increased treatment costs and productivity losses from sickness and premature death. While oxytocin is widely accessible, there are concerns about its quality and the burden of SF oxytocin remains understudied.

**OBJECTIVE:** To develop an impact model to estimate the health and economic burden of SF oxytocin in Kenya. This paper presents the methodology and the findings of assessing SF oxytocin in Kenya.

**METHODS:** A decision tree model was developed to compare health outcomes and costs with and without SF oxytocin from a healthcare sector and societal perspective. This model incorporates healthcare seeking behavior, epidemiological parameters, medicine quality, health outcomes and costs. The main assumption of the model is that lower active pharmaceutical ingredient (API) percentage results in lower medicine efficacy. Sensitivity analyses were performed to evaluate parameter uncertainty.

**FINDINGS:** For 1.1 million pregnant patients delivering in a healthcare facility in Kenya and a 7% prevalence of oxytocin with 75%-90% API, the model estimates that the presence of SF oxytocin in Kenya is associated with 1,484 additional cases of mild PPH, 583 additional cases of severe PPH, 15 hysterectomies, 32 deaths, 633 DALYs accrued, 560 QALYs lost, and 594 years of life lost yearly. The economic burden of SF oxytocin was $1,970,013 USD from a societal perspective, including $1,219,895 from the healthcare sector perspective. Productivity losses included $12,069 due to missed days of work and $725,979 due to premature death.

**CONCLUSIONS:** By providing local estimates on the burden of SF medicines, the model can inform key policy makers on the magnitude of their impact and support initiatives that facilitate greater access to quality medicines.

## 1. INTRODUCTION

Maternal mortality is a major global health concern, especially in low- and middle-income countries (LMICs).(1) Although global maternal mortality declined by 38% between 2000 and 2017, it still remains alarmingly high. Each year, nearly 295,000 women die during and following pregnancy and childbirth with 94% of these deaths occurring in LMICs.(1) In Kenya, the death rate for pregnancy-related causes is 355 per 100,000 live births.(2) This equals approximately 5,000 girls and women dying annually of pregnancy and childbirth complications, most of which could be prevented.(3)

Postpartum hemorrhage (PPH), defined as a blood loss of 500mL or more following childbirth, is a leading direct cause of maternal mortality globally, accounting for 27.1% of all maternal deaths.(4) Over 14 million women experience PPH every year causing 70,000 maternal deaths due to bleeding complications related to pregnancy and childbirth.(5) Women who survive PPH are at subsequent risk for severe maternal morbidities including organ dysfunctions and long-term disabilities.(6,7) When PPH occurs, early identification and management of bleeding using evidence-based guidelines can prevent most PPH-related severe complications and deaths.(8) According to guidelines, the use of uterotonics is key for both the prevention and management of PPH. Specifically, oxytocin is recommended by the World Health Organization (WHO) as the drug of choice for preventing and treating PPH. (9) Oxytocin is included in the WHO Essential Medicines list and in the United Nations Commission on Life-Saving commodities for Women and Children list. (9–11)

While oxytocin is widely accessible, there are concerns about its quality in LMICs. (10,12,13) A systematic review of the literature reported a median prevalence of 45.6% (range 0-80%) of substandard and falsified (SF) oxytocin in LMICs mainly due to inadequate levels of active pharmaceutical ingredients (API) among tested samples. (13) One study conducted in Ghana reported that 74% of oxytocin purchased at private pharmacies did not meet manufacturer specifications for percentage of API.(14) Due to lack of sufficient reporting on medication quality, inconsistencies in study sampling methods and variability in product testing, the estimates of SF oxytocin prevalence have limitations and are likely “just a small fraction of the total problem”.(15)

Research groups have estimated the health and economic impact of selected SF medicines especially antimalarials and antibiotics. However, the burden of SF oxytocin remains an understudied problem.(16,17) A review of specific causes of maternal deaths in Kenya showed that PPH is an issue that needs to be addressed and that over 80% of maternal deaths are attributed to poor quality of care. (18,19) In recognition of the need to combat the threat of SF medicines and to improve the quality of PPH care, we developed a comprehensive disease model to generate country-specific evidence on the health and economic burden of SF oxytocin used for the prevention and treatment of PPH among pregnant women from the first stage of labor until discharge or maternal death. This paper describes the methodology of the model and findings for Kenya.

## 2. MATERIALS AND METHODS

### Model Structure

A decision tree model was developed based on previous SF burden models, economic evaluations of uterotonics in the prevention and treatment of PPH, and a review of clinical practice guidelines. (20–26) The model estimates the health and economic burden of SF oxytocin from a healthcare sector and societal perspective over a lifetime horizon using Microsoft Excel software (Microsoft Corp., Redmond, WA, USA). To estimate the incremental health outcomes and costs of SF oxytocin, our model compares two mutually exclusive scenarios: a real-world scenario based on the estimated prevalence of SF oxytocin and an ideal-world scenario where the prevalence of SF oxytocin is zero.

Figure 1 shows the schematic representation of the model structure and model inputs.

**Figure 1.**
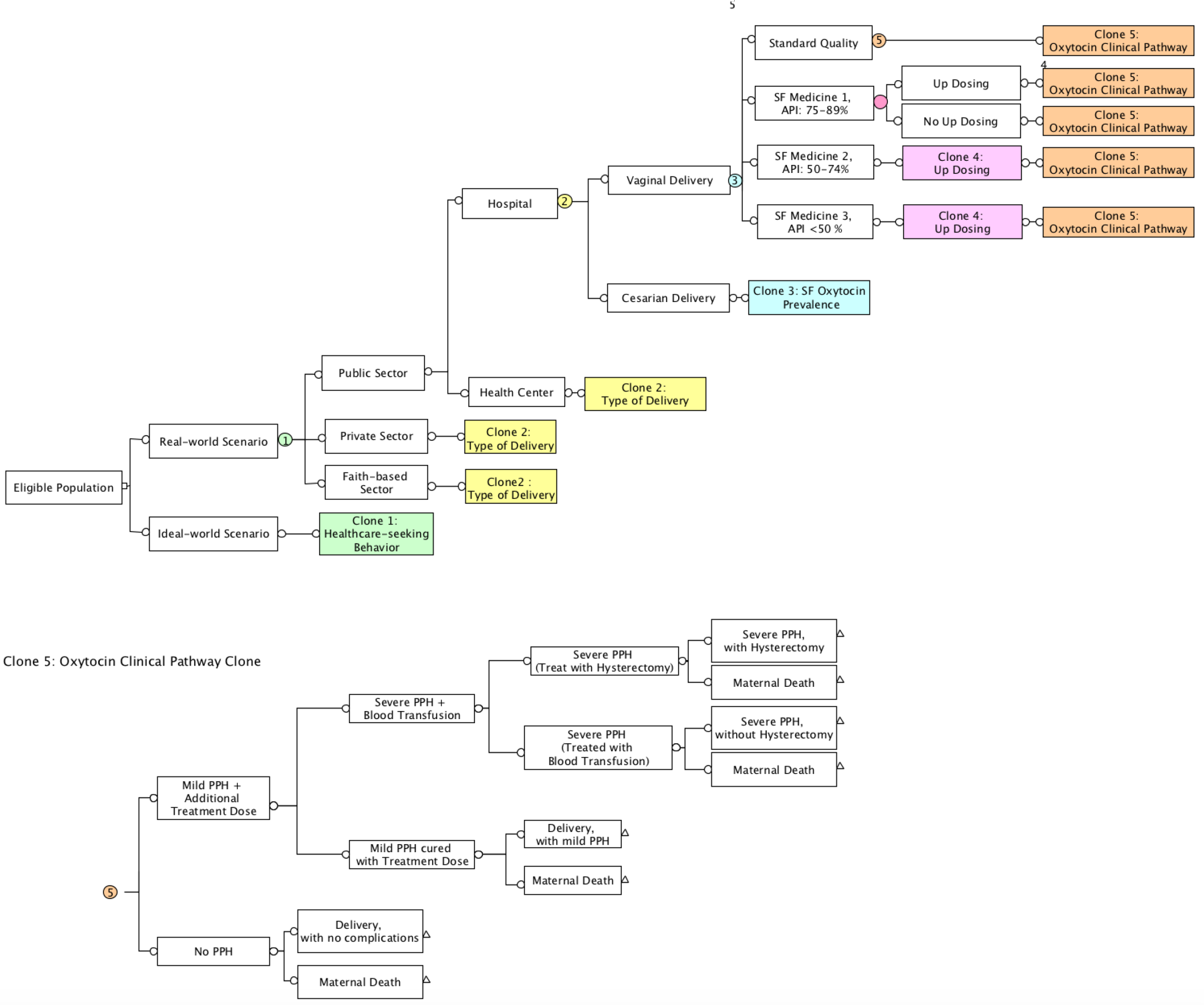
Decision tree model to estimate the health and economic burden of SF oxytocin

A cohort of pregnant women of mean childbearing age, i.e., 28 years old, going into first stage of labor at a healthcare facility enters the model and delivers at a private, public or faith-based healthcare.(27) The cohort of women is divided into two groups: vaginal deliveries and cesarean deliveries. In the model, each group receives a prophylactic dose of oxytocin as recommended by the WHO for PPH prevention. (9)

### Model Inputs

There are four quality categories of oxytocin based on the percentage of the active pharmaceutical ingredient (API):

● Standard quality: 90 -110% of the API as defined by the United States Pharmacopeia (USP) (28)
● Substandard quality and falsified level 1 (SF1): 75 – 89% of the API required by the USP standard (28)
● Substandard quality and falsified level 2 (SF2): 50 – 74% of the API required by the USP standard (21)
● Substandard quality and falsified level 3 (SF3): < 50% of the API required by the USP standard (21)

A proportion of providers are expected to increase the dose (up dose) category SF1, SF2 or SF3 drugs based on a study that found that 41% of the healthcare providers at a healthcare facility in Nigeria doubled the WHO-recommended dose and 10% used two to six vials instead of one to achieve the desired outcome.(29)

After the initial prophylactic dose, a woman can either have an adequate response to the treatment and is discharged or she can experience mild PPH (blood loss <500mL) (9,30) and is given another dose of oxytocin as a first line of PPH treatment following WHO and UK guidelines.(31–33) After a treatment dose of oxytocin, bleeding either stops and the woman is discharged, or the woman does not respond to the treatment and severe PPH persists, in which case, she typically receives intrauterine balloon tamponade, exploration of uterine cavity, uterine compression sutures and a blood transfusion.(34,35) After treatment for persistent severe PPH, the bleeding stops and the woman is discharged; or the treatment fails and a hysterectomy is performed.(35) After the hysterectomy, the two outcomes modeled are either maternal survival with secondary infertility or maternal death.

Each branch in the pathway is associated with costs and health outcomes, which are accumulated over the time horizon of the model, i.e., lifetime, and are discounted at 3%. Incremental cases of mild PPH, severe PPH, hysterectomies and deaths were obtained by multiplying branch probabilities for each health outcome for the real-world scenario and for the ideal-world scenario and the difference was calculated.

The decision tree incorporates the following health states:

1- Delivery, with no complications
2- Delivery, with mild PPH
3- Severe PPH without hysterectomy
4- Severe PPH with hysterectomy
5- Maternal death

Each of these health states was associated with a utility and a disability weight. Utility weights were obtained from a cost-effectiveness study comparing oxytocin and carbetocin where EQ-5D-5L was used to collect primary data on vaginal and cesarian deliveries with and without PPH that was then converted to utility scores.(36) Utility weights for hysterectomy and secondary infertility were obtained from cost-effectiveness studies examining the treatment of heavy menstrual bleeding and menorrhagia.(37,38) Based on the Briones et al. economic evaluation for PPH prophylaxis, we assumed that delivery with no complications, delivery with mild PPH and severe PPH without hysterectomy and severe PPH with hysterectomy lasted 6 weeks. After the six weeks, we assumed that the utility weight retuned to 1, i.e., “perfect health”, for all the aforementioned health states, except for severe PPH with hysterectomies, where the utility weight for secondary infertility was assigned for the remaining of the woman’s lifetime.(38)

We estimated YLLs associated with each health state then and QALYs and DALYs were calculated by multiplying the number of years that the women were expected to be alive (converted from the six-week cycle length used in the analysis) by utility scores and disability weights, respectively.

Six categories of model inputs were included: health-seeking behavior, epidemiological parameters, medicine quality, treatment efficacy, outcome measures and costs. Health-seeking behavior inputs consist of the percentage of women who have a healthcare facility delivery in the private, public or faith-based sector. Epidemiological parameters include the annual number of pregnant women delivering in healthcare facilities, percentage of hospitals and health centers and/or clinics in the public and private sector, probability of hysterectomy if severe PPH occurs, probability of PPH-related mortality and probability of dying by age (using life tables). Medicine quality inputs include the percentage of standard quality medicines and percentage of SF medicines depending on their levels of API. Treatment efficacy included oxytocin treatment effects, probabilities of mild and severe PPH with and without oxytocin, and outcomes measures included health state utilities and disability weights.

We gathered these inputs from peer-reviewed literature, public institutions, recent Kenya Demographics and Health Surveys data (DHS) and secondary data collection from in-country stakeholders in Kenya.(2) The secondary data was obtained through Kenyan subject matter experts in the field of medicine quality and maternal health. The prevalence of SF medicines specifically was the best available estimate provided by them; however, it is based on a small sample size for the entire country, which makes the input highly uncertain.

As for cost inputs, using healthcare sector and societal perspectives, direct and indirect costs were considered in the analysis. The direct medical costs included were drug costs, labor costs (corresponding to time spent by healthcare providers) and hospital costs associated with hospitalizations, operating units, blood transfusions, invasive procedures, and hysterectomy costs. Direct medical costs were either derived from the literature or secondary data collection from a Kenyan technical working group. Indirect costs consisted of productivity losses incurred by patients due to days missed at work or premature maternal death. Productivity losses were estimated using the human capital approach based on gross domestic product (GDP) per capita, duration of lost productivity, and rate of employment. Productivity losses due to premature death were also estimated using the human capital approach, life of years lost and employment rate.(39) All monetary units were converted from Kenyan Shillings (KES) to US Dollars (USD) using the average annual exchange rate for 2022 (1 USD = 122.935 KES) and were rounded to the nearest thousands. (40) Base case inputs were identified as the “most likely” estimates for each data input. For each parameter, we also identified an uncertainty range to capture potential variation from base case values. Uncertainty ranges were either formal estimates of variance when available (e.g., 95% confidence intervals), an evidence-based assumption or a ±10% variation from the base case.

All model inputs, low and high range estimates and distributions are shown in Table 1.

**Table 1.** Model inputs: base-case values, uncertainty ranges, distributional forms and sources

| Parameters | Base-case Value | Uncertainty range (low, high) | Distribution | Sources |
| --- | --- | --- | --- | --- |
| *Population and health-seeking behavior in Kenya* |  |  |  |  |
| Eligible population: Number of pregnant people at | 1,599,306 | (1,507,450, | Normal | (41) |
| third stage of labor |  | 1,723,692) |  |  |
| Percentage of healthcare facility delivery | 75% | (67, 79) | Beta | (41) |
| Percentage of home delivery | 25% | (21, 33) | Beta | (41) |
| Percentage of pregnant people who deliver in the public sector | 70% | (69, 74) | Beta | (41) |
| Percentage of pregnant people who deliver in the private sector, i.e., private hospital | 17% | (15, 18) | Beta | (41) |
| Percentage of pregnant people who deliver in “Other” sector, if any | 13% | (11, 14) | Beta | (41) |
| Proportion of Hospitals in Public Sector | 40% | (36,44) | Beta | (42) |
| Proportion of Public Health Centers in Public Sector | 60% | (54,66) | Beta | (42) |
| Percentage of people who have vaginal delivery | 83.6% | (81, 97.9) | Beta | (41) |
| Percentage of people who have cesarian delivery | 16.4% | (2.1, 19) | Beta | (41) |
| Percentage of healthcare providers who up dose oxytocin | 41% | (45.9, 56.1) | Beta |  |
| Number of doses used to compensate for SF oxytocin during up dosing approach |  |  |  |  |
| SF medicines with 75-89% API | 2 | (1, 3) | Normal | (29) |
| SF medicines with 50-74% API | 3 | (1, 5) | Normal | (29) |
| SF medicines with less than 50% API | 6 | (1, 6) | Normal | (29) |
| Medicine Quality (National Average) |  |  |  |  |
| Prevalence of standard quality medicines (API 90% - 110%) | 93% | (86, 100) | Beta | (43) |
| Prevalence of SF1 medicines (API $\geq 75$ - <90%) | 7% | (0, 14) | Beta | (43) |
| Prevalence of SF2 medicines (API $\geq 50$- <75%) | 0% | N/A | Beta | (43) |
| Prevalence of SF3 medicines (API <50%) | 0% | N/A | Beta | (43) |
| Model assumption:<br>SF medicine efficacy reduction |  |  |  |  |
| Efficacy reduction (75–90% API) | 30% | (15, 45)**Error! Bookmark not defined.** | Beta | (21) |
| Efficacy reduction (50–75% API) | 60% | (45, 75)**Error! Bookmark not defined.** | Beta | (21) |
| Efficacy reduction <50% | 100% | N/A | Beta | (21) |
| Health outcomes |  |  |  |  |
| Vaginal delivery: Probability parameters |  |  |  |  |
| Probability of mild PPH occurrence /needing additional uterotronics with oxytocin | 0.122 | (0.089, 0.172) | Beta | (44) |
| Probability of mild PPH occurrence/needing additional uterotronics without oxytocin | 0.239 | (0.174, 0.337)**Error!** | Beta | (44) |
|  |  | **Bookmark not defined.** |  |  |
| Probability of severe PPH occurrence / needing transfusion with oxytocin | 0.029 | (0.02, 0.041) | Beta | (44) |
| Probability of severe PPH occurrence /needing transfusion without oxytocin | 0.048 | (0.033,0.069) | Beta | (44) |
| Probability of hysterectomy if severe PPH | 0.016 | (0.144, 0.0176) | Beta | (45) |
| Probability of maternal death if hysterectomy | 0.00292 | (0.0006, 0.00378) | Beta | (46) |
| Cesarian delivery |  |  |  |  |
| Probability of mild PPH occurrence with oxytocin | 0.442 | (0.247, 0.791) | Beta | (47) |
| Probability of mild PPH occurrence without treatment | 0.7 | (0.39, 0.8) | Beta | (48) |
| Probability of severe PPH occurrence / needing additional uterotonics with oxytocin | 0.374 | (0.272, 0.524) | Beta | (47) |
| Probability of severe PPH occurrence / needing additional uterotonics without oxytocin | 0.598 | (0.4352, 0.8384) | Beta | (48) |
| Probability of hysterectomy if severe PPH | 0.027 | (0.0243, 0.0297) | Beta | (49) |
| Probability of maternal deaths if severe PPH/hysterectomy | 0.054 | (0.021, 0.13) | Beta | (50,51) |
| Vaginal and C-section deliveries |  |  |  |  |
| Numbers of days missed at work due to mild PPH | 2.98 | (2.94, 3.08) | Normal | (52) |
| Number of days missed at work due to severe PPH | 3.67 | (3.56, 3.78) | Normal | (52) |
| Numbers of days missed at work due to hysterectomy | 7.8 | (0.4, 28) | Normal | (53,54) |
| Quality of life parameters |  |  |  |  |
| Utility weight in vaginal delivery, no complication (6 weeks) | 0.85 | N/A | Beta | (36) |
| Utility weight, mild PPH episode (7 days) | 0.59 | N/A | Beta | (55) |
| Utility weight, severe PPH episode (21 days) | 0.47 | N/A |  | (55) |
| Utility weight in cesarian delivery, no complications (6 weeks) | 0.83 | N/A | Beta | (55) |
| Utility weight hysterectomy, short term (21 days) | 0.42 | N/A | Beta | (55) |
| Utility weight hysterectomy, long term | 0.88 | N/A | Beta | (38) |
| Disability weight in vaginal delivery, no complication | 0.002 | N/A | Gamma | (55) |
| Disability weight for maternal hemorrhage (less than 1L blood loss) | 0.114 | (0.078, 0.159) | Gamma | (56) |
| Disability weight for maternal hemorrhage | 0.324 | (0.220, 0.442) | Gamma | (56) |
| (Greater than 1L blood loss) |  |  |  |  |
| Disability weight hysterectomy | 0.274 | N/A | Gamma | (55) |
| Disability weight in cesarian delivery, no complication | 0.004 | N/A | Gamma | (55) |
| Disability weight of secondary infertility after hysterectomy (Proxy weight of infertility due to puerperal sepsis) | 0.005 | (0.002, 0.011) | Gamma | (56) |
| Utility weight in vaginal delivery, with PPH episode | 0.59 | 0.03 | Beta | (36) |
| Utility weight in cesarian delivery, no complications | 0.47 | 0.02 | Beta | (36) |
| Utility weight in cesarian delivery, with PPH episode | 0.83 | 0.15 | Beta | (36) |
| Utility weight hysterectomy, short term | 0.42 | (0.49, 0.74) | Beta | (37) |
| Utility weight hysterectomy, long term | 0.88 | (0.75, 0.95) | Beta | (38) |
| Disability weight in vaginal delivery, no complication | 0.41 | (0.219, 0.451) | Gamma | (55) |
| Disability weight for maternal hemorrhage (less than 1L blood loss) | 0.114 | (0.078, 0.159) | Gamma | (56) |
| Disability weight for maternal hemorrhage (Greater than 1L blood loss) | 0.324 | (0.220, 0.442) | Gamma | (56) |
| Disability weight hysterectomy | 0.58 | (0.522, 0.638) | Gamma | (55) |
| Disability weight in cesarian delivery, no complication | 0.58 | (0.522, 0.638) | Gamma | (55) |
| Disability weight of secondary infertility after hysterectomy (Proxy weight of infertility due to puerperal sepsis) | 0.005 | (0.002, 0.011) | Gamma | (56) |
| Population characteristics |  |  |  |  |
| Mother's mean childbearing age | 28.6 | (14, 50) | Normal | (27,57) |
| Employment Rate | 49.9% | (29.4, 61.6) | Beta | (58) |
| Costs (\$) | | | | |
| Cost of oxytocin per treatment (\$), national average | 0.39 | (0.03-1.35) | Gamma | (59–61) |
| Cost of oxytocin per treatment (\$), public sector | 0.029 | (0.0232, 0.0348) | Gamma | (61) |
| Cost of oxytocin per treatment (\$), private sector | 1.354 | (1.0832, 1.6248) | Gamma | (61) |
| Cost of oxytocin per treatment (\$), faith-based sector | 0.672 | (0.16-1.354) | Gamma | (60) |
| Cost of additional oxytocin and mild PPH management (Public sector) | 77.61 | (50-500) | Gamma | (34,59) |
| Cost of additional oxytocin and mild PPH management (Private sector) | 400 | (320, 480) | Gamma | (34,59) |
| Cost of additional oxytocin and mild PPH management (Faith-based sector) | 238.8 | (77.61, 400) | Gamma | (34,59) |
| Cost of Cost of severe PPH management (public) | 1,343 | (77.61-1,611) | Gamma | (34,59) |
| Cost of Cost of severe PPH management (private) | 2,686 | (77.61-3,223) | Gamma | (34,59) |
| Cost of Cost of severe PPH management (faith based) | 2,014 | (77.61-2,417) | Gamma | (34,59,62) |
| Cost of hysterectomy (public) | 662 | (640, 2,074)**Error! Bookmark not defined.** | Gamma | (59,63) |
| Cost of hysterectomy (private) | 2,074 | (1,659, 2,489)**Error! Bookmark not defined.** | Gamma | (34) |
| Cost of hysterectomy (faith based) | 1,368 | (1,094, 1,642)**Error! Bookmark not defined.** | Gamma | (59,62) |
| Discount rate | 3% | (0, 5) |  | (64) |

### Model Outcomes

The primary outputs of the model are estimates of the health and economic burden based on the increased morbidity and mortality due to SF oxytocin.

#### a. Health outcomes

Health outcomes estimated by the model are incremental cases of mild PPH, severe PPH, hysterectomies, deaths, years of life lost (YLLs), additional DALYs incurred and QALYs los due to the presence of SF oxytocin.

#### b. Economic outcomes

Economic outcomes in the model are estimated from a healthcare sector and societal perspective and included total costs for the real-world scenario with presence of SF oxytocin, total costs for the ideal-world scenario without SF oxytocin and incremental costs due to the presence of SF oxytocin.

### Assumptions

The main assumption in this model is the relationship between the API percentage and medicine efficacy, based on a burden model developed by Bath et al. (21) This assumption states that standard quality medicines have 100% medicine efficacy. Medicines in the SF level 1 category with API between 75 and 89%, have a 30% efficacy reduction. Medicines in the SF 2 category with API between 50 and 74%, have a 60% efficacy reduction and medicines in the SF 3 category with API less than 50%, have 100% efficacy reduction, i.e., equivalent to not receiving treatment. Additionally, we assumed that oxytocin was the only uterotonic used for the prevention and management of PPH and that the background risk factors for PPH were comparable among women in the real-world scenario and ideal-world scenario.

### Sensitivity analyses

To quantify the impact of parametric uncertainty on model results, a one-way sensitivity analysis (OWSA) and a probabilistic sensitivity analysis (PSA) were performed.

For each outcome, an OWSA assessed the influence of uncertainty in individual model parameters on model results. This was achieved by varying each fixed parameter around the maximum and minimum of its uncertainty interval and plotting the model results in a Tornado diagram.

The PSA was conducted to evaluate the range of plausible outcomes given joint uncertainty in all inputs. We used Monte Carlo simulations with 10,000 iterations, where inputs were randomly and simultaneously generated within a specific distribution.(65) Epidemiological inputs were assumed to be normally distributed. Probabilities and percentages were assumed to have a beta distribution for binomial data inputs or a Dirichlet distribution for multinomial data inputs. Cost inputs were assumed to have a gamma distribution.

### Model Validation

A model validation was performed to evaluate how well the model represents the health and economic burden of SF oxytocin and whether the results of the analysis can serve as a basis for decision making. (66) First, we verified that the model was parsimonious and sufficient to answer the research question by incorporating the main assumptions about SF medicines and the current care of disease progression and treatment strategies.(67) Because there are no other model that examined SF oxytocin specifically, this was performed by comparing our model characteristics and structure to existing models and approaches that estimated the impact of SF medicines and economic evaluations of different uterotonics for the prophylaxis and treatment of PPH during third stage of labor.

We assessed internal validity and consistency by checking that the mathematical equations, calculations and code were accurate and consistent with the specifications of the model.

We performed external validation and consistency, whereby we compared the model outputs for the real-world scenario with current in-country maternal outcomes to ensure that the model adequately reflects the current situation in Kenya. This was performed by comparing the existing estimates of number of PPH episodes, hysterectomies and PPH-related deaths in Kenya as a benchmark for our model predictions.

Three technical working group meetings were held with key stakeholders from Kenya between January and December 2021 to gather subject matter experts’ opinion on data inputs for the model and assumptions. All data inputs and uncertainty ranges were reviewed and discussed by local experts and policymakers from public health and public policy agencies and universities to reach consensus on the most locally representative and recent data to be used in the model.

## 3. RESULTS

### Deterministic base case results

The health and economic outcomes for the real-world and ideal-world scenario for SF oxytocin burden are presented in Tables 2 and 3. For an annual cohort of pregnant women and a prevalence of 7% SF oxytocin, the model estimates that the presence of SF oxytocin in Kenya is associated with 1,484 additional cases of mild PPH, 583 additional cases of severe PPH, 15 hysterectomies, 32 deaths, 633 DALYs accrued, 560 QALYs lost, and 594 years of life lost per year compared to the ideal-world scenario without SF oxytocin.

**Table 2.** Results describing the health burden of SF oxytocin in Kenya for an annual cohort of pregnant women

|  | **Life Years**<br>**(discounted)** | **QALYs**<br>**(discounted)** | **DALYs**<br>**(discounted)** | **Deaths** | **Mild PPH** | **Severe PPH** | **Hysterectomies** |
| --- | --- | --- | --- | --- | --- | --- | --- |
| **Presence of SF** | 22,601,637 | 18,983,693 | 14,766,130 | 1,931 | 174,703 | 35,714 | 950 |
| **No SF** | 22,602,231 | 18,984,253 | 14,765,497 | 1,900 | 173,219 | 35,132 | 935 |
| **Increment** | (594) | (560) | 633 | 32 | 1,484 | 583 | 15 |

**Table 3.**
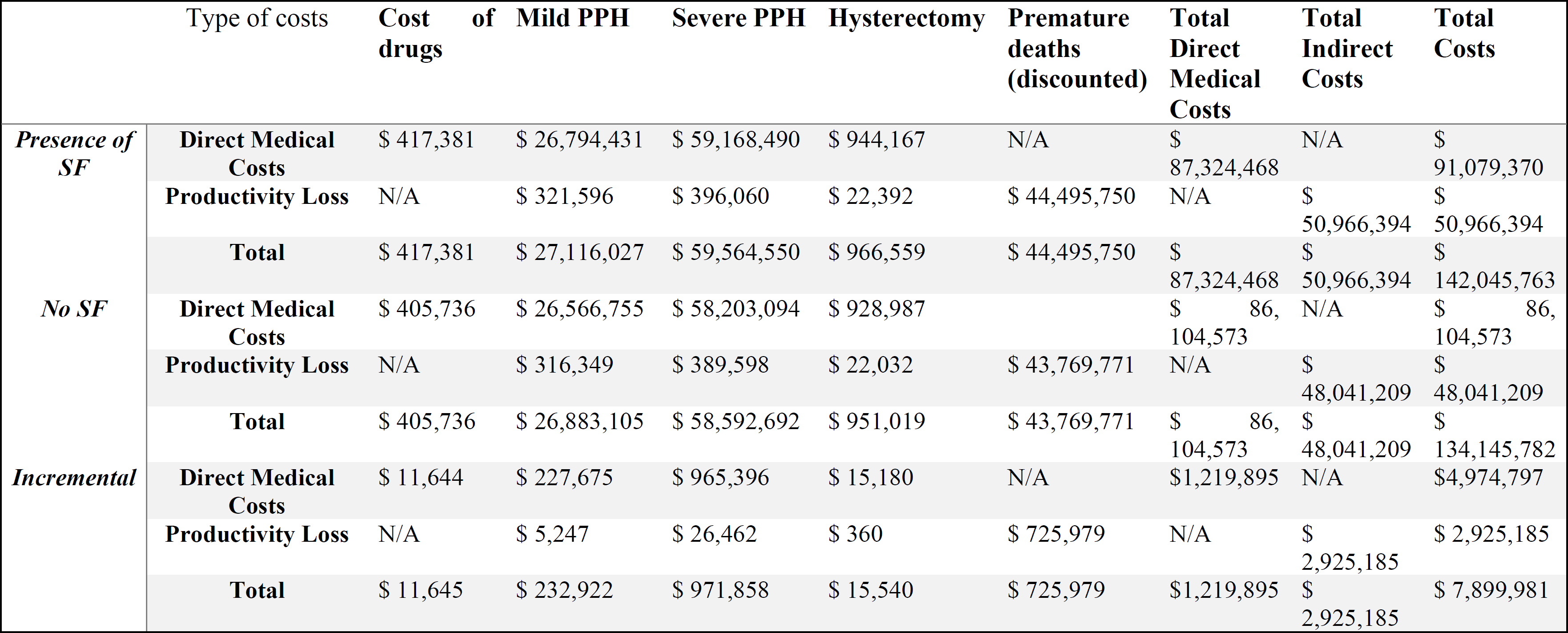
Results describing the economic burden of SF oxytocin in Kenya for an annual cohort of pregnant women

|  | Type of costs | Cost of drugs | Mild PPH | Severe PPH | Hysterectomy | Premature deaths (discounted) | Total Direct Medical Costs | Total Indirect Costs | Total Costs |
| --- | --- | --- | --- | --- | --- | --- | --- | --- | --- |
| **Presence of SF** | **Direct Medical Costs** | \$ 417,381 | \$ 26,794,431 | \$ 59,168,490 | \$ 944,167 | N/A | \$ 87,324,468 | N/A | \$ 91,079,370 |
| | **Productivity Loss** | N/A | \$ 321,596 | \$ 396,060 | \$ 22,392 | \$ 44,495,750 | N/A | \$ 50,966,394 | \$ 50,966,394 |
| | **Total** | \$ 417,381 | \$ 27,116,027 | \$ 59,564,550 | \$ 966,559 | \$ 44,495,750 | \$ 87,324,468 | \$ 50,966,394 | \$ 142,045,763 |
| **No SF** | **Direct Medical Costs** | \$ 405,736 | \$ 26,566,755 | \$ 58,203,094 | \$ 928,987 | | \$ 86,104,573 | N/A | \$ 86,104,573 |
| | **Productivity Loss** | N/A | \$ 316,349 | \$ 389,598 | \$ 22,032 | \$ 43,769,771 | N/A | \$ 48,041,209 | \$ 48,041,209 |
| | **Total** | \$ 405,736 | \$ 26,883,105 | \$ 58,592,692 | \$ 951,019 | \$ 43,769,771 | \$ 86,104,573 | \$ 48,041,209 | \$ 134,145,782 |
| **Incremental** | **Direct Medical Costs** | \$ 11,644 | \$ 227,675 | \$ 965,396 | \$ 15,180 | N/A | \$1,219,895 | N/A | \$4,974,797 |
| | **Productivity Loss** | N/A | \$ 5,247 | \$ 26,462 | \$ 360 | \$ 725,979 | N/A | \$ 2,925,185 | \$ 2,925,185 |
| | **Total** | \$ 11,645 | \$ 232,922 | \$ 971,858 | \$ 15,540 | \$ 725,979 | \$1,219,895 | \$ 2,925,185 | \$ 7,899,981 |

In addition to the health burden, these health outcomes contributed to a total estimated economic burden of $1,970,013 USD in Kenya from a societal perspective over a lifetime horizon. Approximately 37% of the economic burden ($738,049) was due to productivity losses. These productivity losses included $12,069 due to missed days at work as a result of care seeking and convalescence, and $725,979 economic losses due to premature death. The remaining 63% of the economic burden ($1,219,895) was due to direct medical costs from a healthcare sector perspective. The direct medical costs included drug costs ($11,645), mild PPH management costs ($227,675), severe PPH management costs ($965,395) and hysterectomy management costs ($15,180). Costs and health outcomes occurring in the first 6 weeks were not discounted. QALYs, DALYs, life years and costs due to productivity loss due to premature death were discounted at 3%

### Sensitivity Analysis

#### One-way sensitivity analysis

The impact of a one-way sensitivity analysis on incremental societal costs are presented in a tornado diagram (Figure 2). The analysis found that the most sensitive variables were the proportion of SF 1 in the real-world scenario followed by PPH-related mortality and treatment effect of SF 1, i.e., efficacy reduction. s

**Figure 2.**
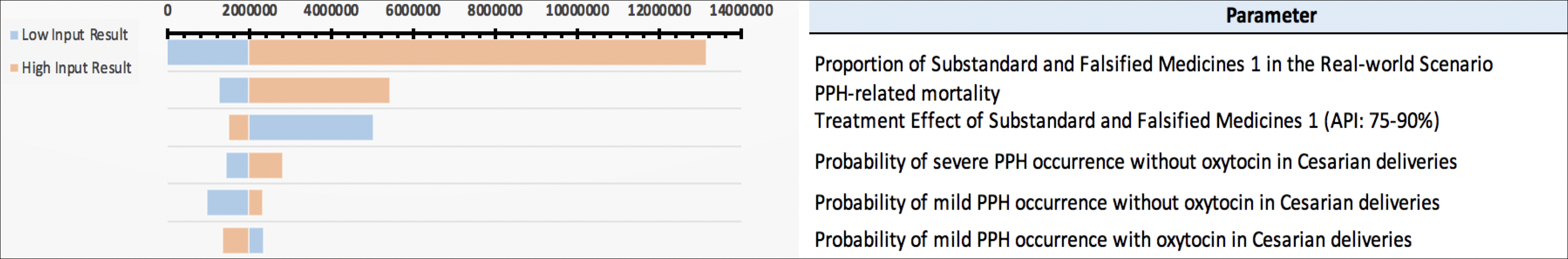
Tornado Diagram for the total costs due to SF oxytocin using oxytocin-specific model

In the event of higher prevalence of SF oxytocin with API between 75% and 89% (SF1) that we used in our base case analysis, significant increases in morbidity and costs would occur. With 14% prevalence of SF1 medicine, SF oxytocin can be associated with up to an additional 15,094 cases of mild PPH, 3,625 cases of severe PPH, 96 cases of hysterectomies and 196 deaths compared to base case results where we assumed 7% prevalence. The economic burden can also vary between $0 and $ 13,133,145 USD from a societal perspective.

#### Probabilistic sensitivity analysis

A probabilistic sensitivity analysis (PSA) was also performed and the results of 10,000 runs are plotted in Figure 3 for societal costs. We notice that 65% of the simulations are below $2M and 90% of simulations are below $4M. Across all simulations, SF oxytocin was found to be associated with a mean societal cost of $2M and resulted in a mean increase of 1,482 mild PPH cases, 596 severe PPH cases, 16 hysterectomies and 33 deaths.

**Figure 3.**
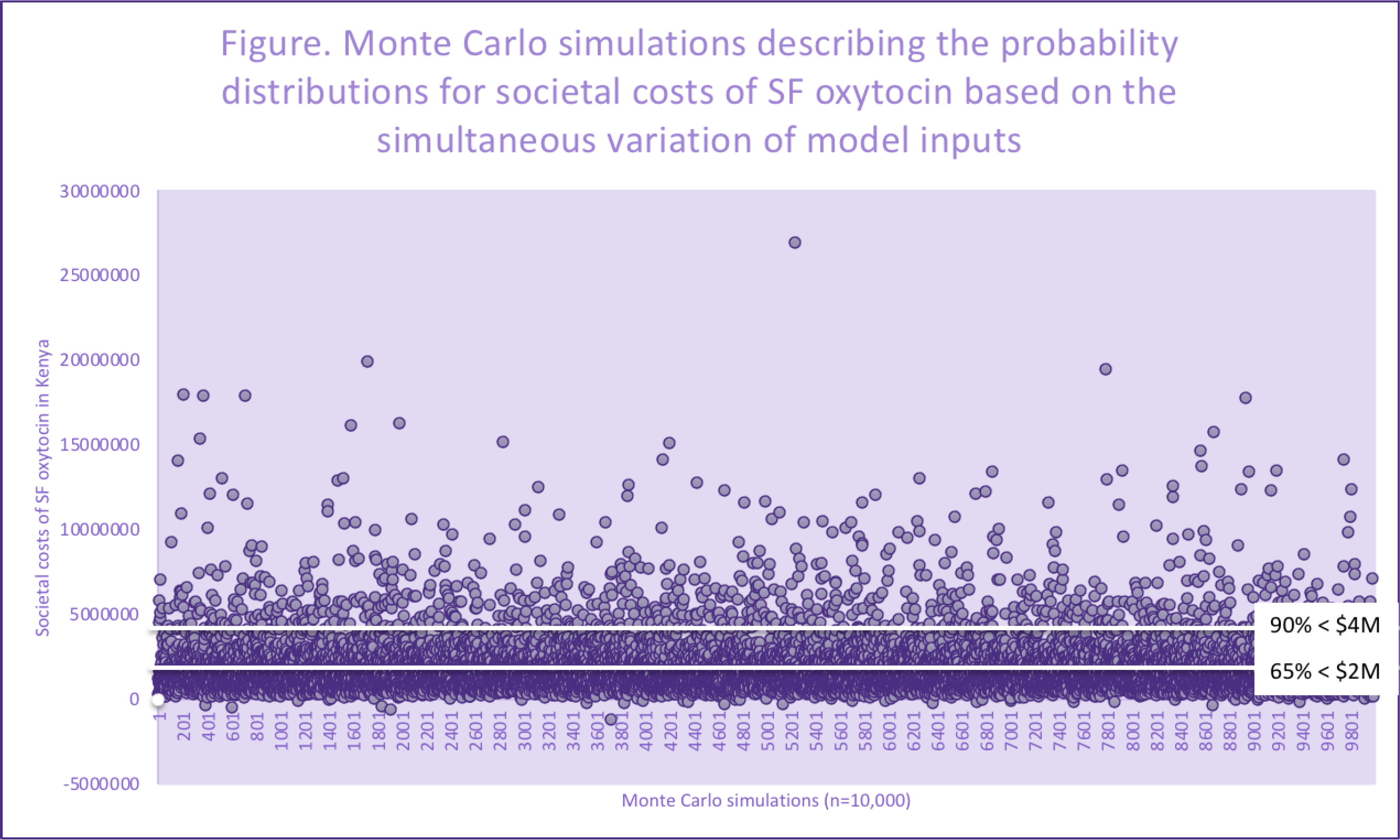
Monte Carlo simulations describing the probability distributions for societal costs of SF oxytocin based on the simultaneous variation of model inputs

## 4. DISCUSSION

The decision tree model can be used to estimate the health and economic burden of SF oxytocin in Kenya from the healthcare sector and societal perspectives. The model follows a cohort of pregnant women of mean childbearing age in a real-world scenario where SF oxytocin is compared to an ideal-world scenario without SF oxytocin. In both scenarios, the model incorporates women’s care-seeking behaviors, their mode of delivery, the progression from first to third stage of labor and its management, the treatment outcomes, and associated costs.

Our findings indicate that every year, the presence of 7% SF oxytocin with API between 75% and 89% in Kenya contributes to the burden of PPH, the leading cause of maternal mortality in the country, with a total of 10,580 episodes of severe bleeding due to SF oxytocin under base case assumptions. A recent study using the Kenyan Confidential Enquiry into Maternal Deaths (CEMD), a database that includes all maternal deaths reported through the District Health Information system (DHIS2), recently estimated that between 2014 and 2017, 728 deaths of a total of 2,292 maternal deaths were due to PPH, or an average of 182 PPH-related death, annually.(68) Data frames and time periods are different, so comparison should be cautious, but contrasting these findings with our model estimates of 105 deaths due to PPH per year indicates that SF oxytocin may be contributing significantly to annual PPH-related maternal mortality in Kenya. (68) That is particularly important as the 7% prevalence of SF is highly uncertain and may not be representative of exposures across the country. This input was based on the Kenyan technical group’s expertise and not empirical studies. The samples collected for medicine quality assessments by the Pharmacy and Poison Board are typically small (n=100s) for the entire country and across many different pharmaceutical categories. Because our findings are based on the assumed prevalence of 7% SF oxytocin, it is important to interpret the findings of this study cautiously. Uncertainty was incorporated in the OWSA with a low value of 0% and a high value of 14%, and SF oxytocin prevalence was found to have the most effect on the health and economic burden of SF oxytocin which varies drastically if the prevalence was significantly higher or lower. The economic burden of SF oxytocin is also substantial as our model estimates that almost USD $8 million are expended on direct and indirect costs every year due to SF medications. In our analysis, the main cost drivers are the management of severe PPH, followed by productivity losses due to premature deaths. Despite the common practice of updosing in Kenya, our model estimated that drug costs constituted only 0.5% of the total cost of PPH management. These findings align with a study conducted in the United Kingdom that found that drug costs also only accounted for 0.5% of the total cost of PPH management, suggesting that the bulk of direct costs is allocated to medical procedures and to staff labor necessary to stop severe PPH. (69)

Our model builds upon previous efforts by Bath et al. and Ozawa et al. that estimated the health and economic burden of antimalarials and antibiotics. (21,70) For example, the efficacy reduction assumption utilized in these models are comparable to the assumptions we use. However, there are several limitations in our model. Our model uses decision tree modeling and cannot account for national, regional and patient-specific variations, i.e., heterogeneity within the cohort of pregnant women. However, we can change the mean age and the number of pregnancies of a cohort. Several studies found that hypertension, prolonged labor, advanced maternal age, multiple pregnancies were among the most common risk factors for PPH. (71–73) Because the same population is entering the model in two different scenarios, i.e., real-world and ideal-world scenario, the distribution of risk factors for PPH among women is similar; therefore, these factors are not accounted for in the model. Another limitation of our analysis is that we assumed that oxytocin was the only uterotonic used for the management of third stage labor, as it is WHO’s recommendation for the prevention and management of PPH. We are not accounting for women who may be receiving other uterotonics such as misoprostol, carbetocin or ergometrin. With this approach, we may be overestimating the cost of maternal healthcare as oxytocin is more expensive to acquire, store and handle than some of the other uterotonics used for the same purpose. Oxytocin may also not always be the best option especially for women with pre-existing conditions or complications such as hypertension, heart disease or allergies so we may also be overestimating maternal health outcomes. Lastly, the quality of the data inputs inherently limits the findings of our model. We performed an extensive literature search to ensure the most recent and highest quality peer-reviewed data were used. We also held three meetings with subject matter experts from Kenya to validate all data inputs and their ranges to ensure local relevance and performed a OWSA and a PSA with wide ranges to address uncertainty. Our oxytocin-specific model can be used in Kenya and other countries where maternal mortality is high to provide country-specific estimates of avertable health outcomes and costs due to SF oxytocin or to evaluate the impact of interventions combatting SF oxytocin. The findings can be leveraged by policy makers to respond with an evidence-based strategy to combat SF oxytocin at the country level.

## 5. CONCLUSIONS

The Kenyan government’s efforts over the last decade to invest in maternal health led to a 30-percent decline from 488 deaths per 100,000 live births in 2010 to 342 deaths per 100,000 live births in 2017 but the maternal mortality ratio still remains high compared to the Sustainable Development Goal to reduce the global maternal mortality ratio to less than 70 per 100,000 live births, with no country having a maternal mortality ratio exceeding twice the global average.(1,74,75) The majority of maternal deaths are due to PPH and can be prevented if access to high quality care and oxytocics during and after childbirth is available. Our study demonstrates that SF oxytocin contributes significantly to the health and economic burden of maternal morbidity and mortality and emphasizes the importance of reducing the health and economic burden of SF oxytocin through providing access to better quality medications.

## Data Availability

All relevant data are within the manuscript and its Supporting Information files.

## Acknowledgements

We thank the members of the technical working group in Kenya led by Edward Abwao, Karim Wanga and Zahida Qureshi for their support with secondary data collection and the contributions from the PQM+ program (Leslie Rider-Araki and Souly Phanouvang) for facilitating country engagement with Kenya.

